# The rapid emergence of *Salmonella* Typhi with decreased ciprofloxacin susceptibility following an increase in ciprofloxacin prescriptions in Blantyre, Malawi

**DOI:** 10.1101/2023.03.27.23287794

**Authors:** Philip M. Ashton, Angeziwa Chunga Chirambo, James E. Meiring, Priyanka D. Patel, Maurice Mbewe, Niza Silungwe, Kenneth Chizani, Happy Banda, Robert S. Heyderman, Zoe A. Dyson, Peter MacPherson, Marc Y.R. Henrion, STRATAA Study Group, Kathryn E. Holt, Melita A. Gordon

**Affiliations:** Malawi-Liverpool Wellcome Programme, Blantyre, Malawi; Institute of Infection, Veterinary & Ecological Sciences, University of Liverpool, Liverpool, United Kingdom; Department of Medical Laboratory Sciences, Kamuzu University of Health Sciences, Blantyre, Malawi; Department of Infection, Immunity and Cardiovascular Disease, University of Sheffield, Sheffield, United Kingdom; Research Department of Infection, Division of Infection and Immunity, University College London, London, United Kingdom; Faculty of Infectious and Tropical Diseases, London School of Hygiene & Tropical Medicine, London WC1E 7HT, United Kingdom; Department of Infectious Diseases, Central Clinical School, Monash University, Melbourne, Victoria 3004, Australia; Wellcome Sanger Institute, Wellcome Genome Campus, Hinxton, Cambridgeshire, United Kingdom; School of Health & Wellbeing, University of Glasgow, Glasgow, United Kingdom; Department of Clinical Sciences, Liverpool School of Tropical Medicine, Liverpool, United Kingdom

## Abstract

Ciprofloxacin is the first-line drug for treating typhoid fever in many high burden countries in Africa, but the emergence of non-susceptibility poses a grave challenge to public health programmes. Through enhanced surveillance as part of vaccine evaluation, we set out to investigate the occurrence and determinants of ciprofloxacin non-susceptibility in Blantyre, Malawi.

We performed systematic typhoid fever and antibiotic prescription surveillance in two health centres in Blantyre, Malawi between 01/10/2016 and 31/10/2019, as part of the STRATAA and TyVAC studies. Blood culture isolates from study participants underwent i) pefloxacin screening and ciprofloxacin E-tests to identify ciprofloxcain non-susceptibility and ii) whole genome sequencing (WGS) to identify drug resistance mutations and phylogenetic relationships between non-susceptible and sensitive isolates. We constructed generalised linear regression models to investigate associations between ciprofloxacin prescription rates and *S.* Typhi isolates with Quinolone Resistance Determining Region (QRDR) mutations.

We carried out 11295 blood cultures and microbiologically confirmed 239 cases of typhoid fever, with isolates from 193 participants sequenced (mean age of participants with sequenced genomes 12.8 years, 47% male). Between October 2016 and August 2019 2% (n=4/175) of WGS-confirmed typhoid fever cases were caused by *S.* Typhi with QRDR mutations, compared with 33% (n=6/18) in September and October 2019. Nine of the ten *S.* Typhi with QRDR mutations had a decreased ciprofloxacin susceptibility phenotype. Every additional prescription of ciprofloxacin given to study participants in the preceding month was associated with a 4.2% increase in the relative risk of isolating *S.* Typhi with a QRDR mutation (95% CI, 1.8-7.0%, p=0.0008). Phylogenetic analysis showed that *S.* Typhi isolates with QRDR mutations in September/October 2019 belonged to two distinct sub-clades encoding two different QRDR mutations, and were closely related (0-6 SNPs) to susceptible *S.* Typhi endemic to Blantyre.

We have shown a close temporal association between empiric antimicrobial usage with an increase of fluoroquinolone non-susceptibility in *S*. Typhi, with two sub-clades responsible for the increase. Decreasing ciprofloxacin usage by improving typhoid diagnostics could help to limit the emergence of resistance.

## Intro

*Salmonella enterica* subspecies *enterica* serovar Typhi is the causative agent of typhoid fever, a systemic infectious disease. Globally, in 2017 there were an estimated 10.9 million cases of typhoid fever (95% confidence interval [CI]: 9.3-12.6 million), resulting in 116,800 deaths (95% CI: 65,400 – 187,700), mainly in countries in South and Southeast Asia and Sub-Saharan Africa (Stanaway et al. 2019).

Drug resistance is a major problem in *S.* Typhi infection, leading to worse outcomes for patients and having substantial implications for overstretched health systems in low-and middle income countries (Kaljee et al. 2018). At least 75% of *S.* Typhi in Africa has been identified to be multidrug-resistant (MDR, with resistance to first-line antibiotics including: ampicillin, chloramphenicol, and co-trimoxazole) (Zaki and Karande 2011). The MDR phenotype was associated with the emergence of the H58 lineage in South and Southeast Asia and East and Southern Africa (Wong et al. 2015; Pragasam et al. 2020; Feasey et al. 2015; Kariuki et al. 2010).

*Salmonella* that are resistant to fluoroquinolones (e.g. ciprofloxacin) are amongst the World Health Organization’s (WHO) priority drug-resistant pathogens (‘WHO Publishes List of Bacteria for Which New Antibiotics Are Urgently Needed’ 2017). Changes in the susceptibility of *S.* Typhi to ciprofloxacin are primarily mediated through one of two mechanisms. Firstly, non-synonymous mutations in the quinolone resistance determining regions (QRDR) of DNA gyrase and DNA topoisomerase IV subunits encoded by genes *gyrA, gyrB, parC* and *parE* (Hopkins, Davies, and Threlfall 2005; Cuypers et al. 2018). Secondly, plasmid mediated quinolone resistance (PMQR) which occurs when an organism acquires a plasmid encoding proteins that provide physical protection from the quinolones; enzymes that modify quinolones; or efflux pumps. In Asia, decreased ciprofloxacin susceptibility (DCS, defined as a ciprofloxacin minimum inhibitory concentration [MIC] >0.06 and <1 µg/µl) *S.* Typhi accounted for more than 60% of typhoid cases in Asia in 2011-15, which has led to the adoption of azithromycin and third generation cephalosporins as the first line therapy for typhoid fever in this region (Britto et al. 2018). In the African region, the proportion of *S.* Typhi cases with DCS is below 20% (Britto et al. 2018), although there is substantial variation between countries (Park et al. 2018; Mutai et al. 2018; Kariuki et al. 2021; Ochieng et al. 2022; Acheampong et al. 2019; Al-Emran et al. 2016; Lunguya et al. 2012; Smith et al. 2010). In Zimbabwe, the proportion of *S.* Typhi referred to the national reference lab that were resistant to ciprofloxacin increased from 4% to 22% between 2012 and 2017 (Mashe et al. 2019), while a large outbreak of typhoid fever in Harare showed ciprofloxacin resistance rates of 33% amongst hospitalised patients (N’cho et al. 2019). WGS showed that the ciprofloxacin non-susceptibility/resistance was due to PMQR, in some cases combined with QRDR mutations (Mashe et al. 2021).

In Malawi, an enhanced passive surveillance study in the Ndirande urban township area of the city of Blantyre found an adjusted typhoid incidence of 444 (95% CI 347-717) cases per 100,000 person-years (Meiring et al. 2021). Ninety-two per cent of the *S.* Typhi isolated in this surveillance study were MDR, but only 0.9% of isolates had DCS (Meiring et al. 2021). These findings are consistent with a previous report from a hospital cohort in which 0/176 DCS *S.* Typhi were identified (Feasey et al. 2015).

Against a background of a low rate of ciprofloxacin non-susceptibility in *S.* Typhi in Malawi, we investigated the genomic and epidemiological features of a cluster of *S.* Typhi with QRDR mutations isolated from Blantyre, Malawi in September and October 2019.

## Methods

### Setting

Participants were enrolled either from the community-based primary health centres in Ndirande or Zingwangwa townships (between October 2016-October 2019) or Queen Elizabeth Central Hospital (QECH, [between January 2019 and January 2022]), in Blantyre, Malawi (Darton et al. 2017; Meiring et al. 2019). Ndirande township is an informal urban settlement with a population of around 100,000 people. Population density is around 15,000 people per km^2^, and approximately 85% of residents drink water from shallow wells, rain water, or boreholes, with small numbers having access to municipal water supplies (Kamanula, Zambasa, and Masamba 2014). Zingwanga is a similar setting to Ndirande. QECH, the largest government hospital in Malawi, has 1350 beds and provides free secondary and tertiary healthcare to the approximately 1.3 million residents of Blantyre District and acts as a tertiary referral centre for the Southern Region of Malawi. It admits approximately 10,000 adult and 25,000 paediatric patients per year.

### Sampling

Participants who contributed bacterial isolates for this study were recruited from two main sources. Firstly, the routine blood culture service provided to QECH by the Malawi-Liverpool-Wellcome Programme (MLW). This service is provided for admitted adults (>16 years old) with axillary temperature over 37.5°C or clinical suspicion of sepsis, and for children (<16 years old) who were malaria slide negative, or malaria slide positive and critically ill, or with clinical suspicion of sepsis. We included all *S*. Typhi isolates obtained by this service from January 2019 to January 2022 for which pefloxacin disk diffusion susceptibility testing results were available.

Secondly, the Strategic Typhoid Alliance Across Africa & Asia (STRATAA) and Typhoid Vaccine Acceleration Consortium (TyVAC) studies, the detailed methods of which have been previously published (Darton et al. 2017; Meiring et al. 2019; 2021). Briefly, individuals who were resident in the demarcated geographical area for the STRATAA study (i.e. Ndriande township) or had been enrolled into the TyVAC trial and received a study vaccine were encouraged to present to one of the study facilities when they developed a fever. The STRATAA study enrolled participants from a demographic census of 102,242 individuals in Ndirande township (Meiring et al. 2021) whilst the TyVAC trial enrolled and vaccinated 28,130 children from within the Ndirande and Zingwangwa townships (Patel et al. 2021). A schematic diagram outlining the relationship between the STRATAA and TyVAC cohorts can be seen in Supplementary Figure 1. Participants who received either the candidate or placebo vaccine from the TyVAC trial, or participants from the censused areas (STRATAA study) who presented to a study passive surveillance site and reported a history of fever for ≥72 hours or had a measured axillary temperature of ≥38.0°C, were recruited by members of the passive surveillance study team.

Participants had clinical data collected along with 3-5 ml of blood for microbiological culture. Microbiological culture of blood from all sites/studies was performed at the MLW laboratory on all collected samples with an automated system (BD BACTEC Blood Culture System [Becton-Dickinson, Franklin Lakes, NJ, USA] or BacT/ALERT [BioMerieux, Marcy-l’Étoile, France]) after collection of a single aerobic bottle. This study includes participants from STRATAA or TyVAC recruited between 01/10/2016 and 31/10/2019. Participants were provided with standard of care clinical management at the health centres, with the additional benefit of receiving a diagnostic blood culture. If required, participants were referred to QECH for inpatient care. If participants were blood-culture positive, they were contacted by the clinical team to ensure they remained well and were on the correct antimicrobial therapy. All enrolled participants had antimicrobial usage for the two weeks prior to enrollment and antimicrobial prescriptions on the day of enrollment recorded in the electronic case record form.

### Microbiology & molecular biology

The pefloxacin disk diffusion method was used to detect isolates with altered susceptibility to fluoroquinolones according to the European Committee on Antimicrobial Susceptibility Testing (EUCAST) guidelines. Isolates were cultured on Mueller-Hinton agar with a 5 µg pefloxacin disk, any isolate with a zone of inhibition less than 24mm was considered to be resistant and the ciprofloxacin phenotype determined by E-test. Ciprofloxacin E-tests were performed on Mueller-Hinton agar according to the manufacturer’s instructions (BioMerieux, Marcy-l’Étoile, France), isolates with an MIC of >0.06-1 µg were considered to have decreased ciprofloxacin susceptibility, isolates with an MIC >1 µg were considered to be resistant (Hassing et al. 2013). Isolates were cultured overnight and genomic DNA was extracted using the Wizard Genomic DNA Extraction Kit following the manufacturer’s recommendations (Promega, WI, USA). Genomic DNA was shipped to the Wellcome Sanger Institute for indexed whole genome sequencing on an Illumina HiSeq 2500 platform to generate paired-end reads of 100 bp in length. A subset of genome sequences from participants enrolled in STRATAA between October 2016 and August 2019 have been reported in the multi-site analysis for that study (Dyson et al. 2023).

### Bioinformatics

Sequencing data were trimmed using bbduk v38.96 (‘BBMap’ n.d.) in order to remove adapters and low-quality sequencing regions ‘bbduk.sh ref=adapters.fa in=R1.fastq in2=R2.fastq out=R1.trimmed.fastq.gz out2=R2.trimmed.fastq.gz ktrim=r k=23 mink=11 hdist=1 tbo tpe qtrim=r trimq=20 minlength=50’. Trimmed FASTQs were analysed with mykrobe v0.10.0 using the docker image ‘flashton/mykrobe_for_typhi:latest’ and the GenoTyphi panel i.e. ‘--species typhì (Bradley et al. 2015; Dyson and Holt 2021) to identify the sub-clade of *S.* Typhi. Antimicrobial resistance genes and mutations were identified using amr-finder-plus v3.9.8 with the’-O Salmonellà option (Feldgarden et al. 2019).

Trimmed FASTQs were also mapped against the CT18 reference genome (NCBI accession AL513382.1) using bwa mem v0.7.17-r1198-dirty (Li 2013). SNPs were called with GATK version 3.8-1-0-gf15c1c3ef in unified genotyper mode (Van der Auwera and O’Connor 2020). Positions where the majority allele accounted for < 90% of reads mapped at that position, which had a genotype quality of <30, depth <5x, or mapping quality <30 were recorded as Ns in further analyses. A consensus genome was generated for each genome. These steps were carried out using the PHEnix pipeline https://github.com/phe-bioinformatics/PHEnix. Maximum likelihood phylogenies were then created using IQ-TREE v1.6.12 with in-built model selection (Minh et al. 2020; Kalyaanamoorthy et al. 2017). Phylogenetic trees were annotated using iTOL (Letunic and Bork 2019).

### Statistical analysis

To investigate the association between the proportion of total sequencing-confirmed typhoid fever cases with QRDR mutations and the total number of ciprofloxacin prescriptions in the preceding month, we constructed a generalised linear regression model with a binomial distribution. Analysis was done in R version 4.1.0 and links to a Github repository containing R code to reproduce the all analyses is available in the Data Availability section of the manuscript.

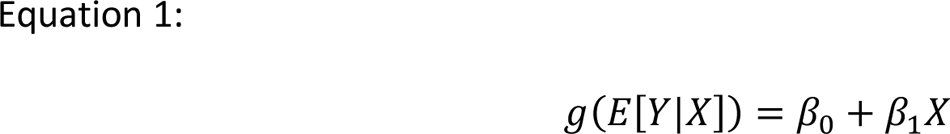

where Y is a binomial random variable recording the QRDR typhoid status for a given sample, X is recording the total number of ciprofloxacin prescriptions in the preceding month and g is the logit function, i.e. 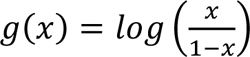

## Results

### Ciprofloxacin susceptibility *S.* Typhi in Blantyre

We screened 503 *S.* Typhi isolated from patients presenting at QECH between January 2019 and January 2022 for resistance to pefloxacin and identified 37 isolates (7%) that were resistant (Figure 1). Following confirmatory ciprofloxacin E-tests, 30 isolates (6% of total) had either decreased ciprofloxacin susceptibility (n = 29) or ciprofloxacin resistance (n = 1).

**Figure 1:**
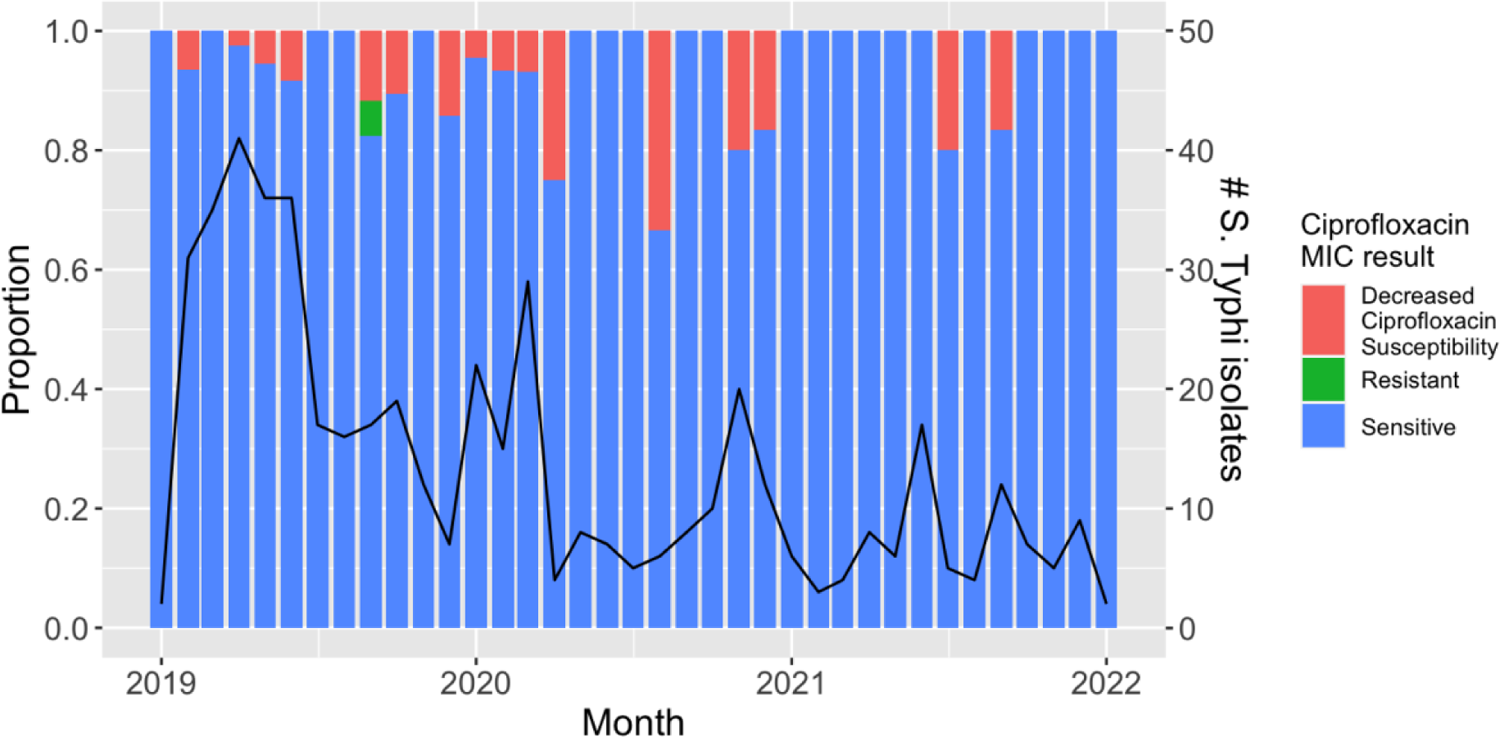
The proportion of *S.* Typhi isolated from QECH that have a sensitive, decreased ciprofloxacin susceptibility, or ciprofloxacin resistant phenotype per month. The total number of *S.* Typhi isolated from QECH is plotted as a black line.

### An increase in the proportion of *S.* Typhi with QRDR mutations was associated with an increase in ciprofloxacin prescriptions

Between 1^st^ October 2016 and 31^st^ October 2019, we carried out enhanced passive surveillance for *S.* Typhi at the Ndirande and Zingwangwa Health Centres, identifying a total of 242 *S.* Typhi isolates from 239 patients (Supplementary Figure 2). Primary results from sequencing of isolates obtained as part of the STRATAA study between October 2016 and August 2019 are reported elsewhere (Dyson et al. 2023), here we report a combined analysis of the 141 Malawian genomes from Dyson et al., 2023 and 55 novel genomes from 52 patients, extending coverage of genomic surveillance to October 2019. In total, from patients recruited between October 2016 and October 2019, we sequenced 196/242 *S.* Typhi from 193/239 patients (Supplementary Figure 2, Supplementary Figure 3) and identified *S.* Typhi from 10 patients with non-synonymous mutations in the quinolone resistance determining regions (QRDR, Figure 2).

**Figure 2:**
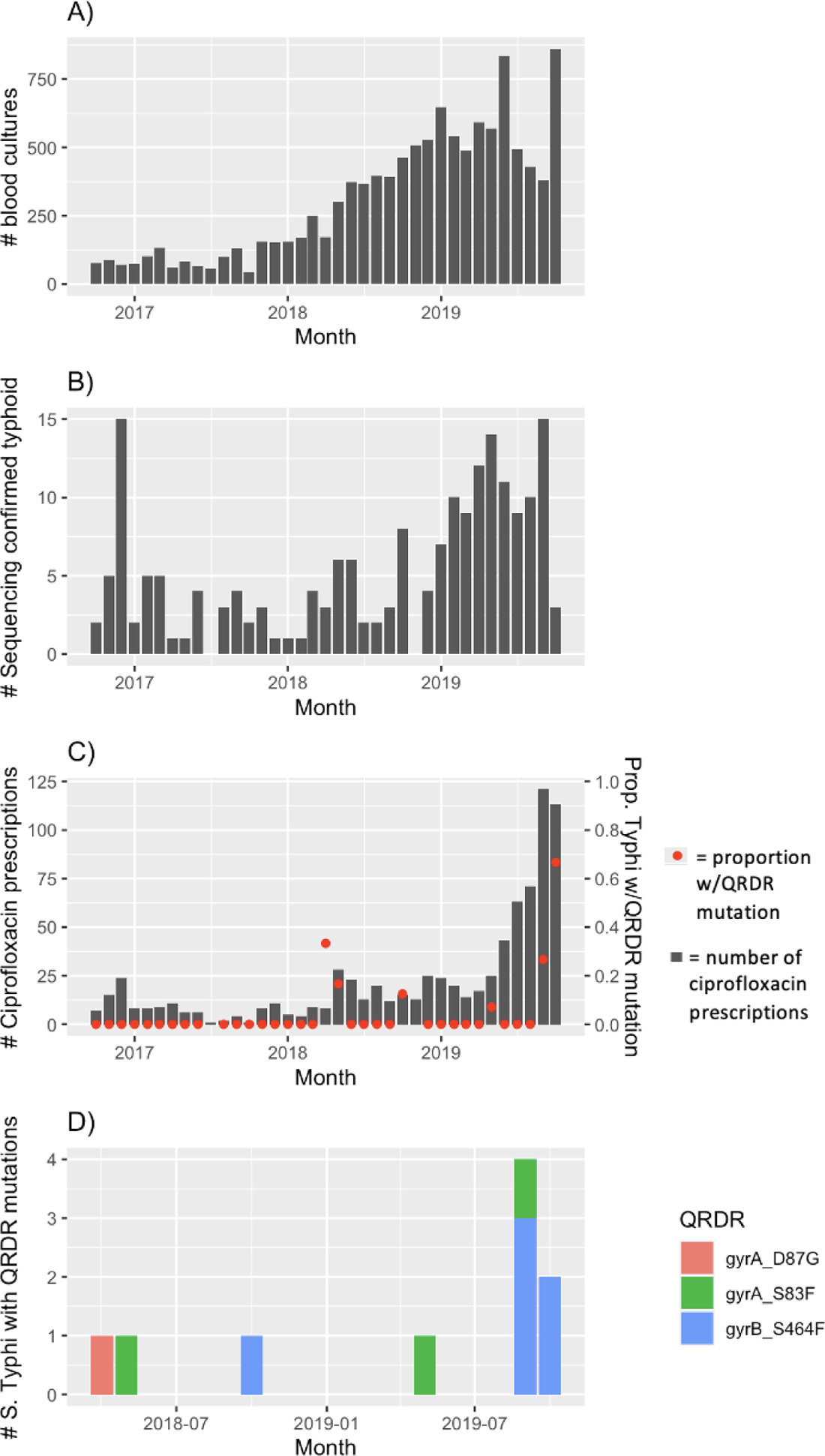
Results of enhanced passive surveillance at Ndirande and Zingwangwa Health Centres A) the total number of blood cultures taken from eligible participants B) Total sequencing confirmed typhoid cases C) the number of ciprofloxacin prescriptions (grey bars) and the proportion of sequencing confirmed *S.* Typhi with QRDR mutations D) the frequency of *S.* Typhi with QRDR mutations, coloured by the QRDR mutation identified.

In the first half of 2019, there was an increase in MDR typhoid fever. Between October 2016 and December 2018 there was a median of three sequencing-confirmed cases of typhoid fever per month, increasing to a median of 10 cases between January and October 2019 (Figure 2B). Following this increase in MDR typhoid, the number of ciprofloxacin prescriptions increased in the second half of 2019, from a median of 11 (range = 1-28) monthly prescriptions between October 2016 and May 2019 to 71 (range = 43-121) monthly prescriptions between June-October 2019 (Figure 2C). Not long after the increase in ciprofloxacin prescriptions, we noted an increase in DCS *S.* Typhi; between October 2016 and August 2019, 4/175 (2%) cases of typhoid fever were caused by *S.* Typhi with QRDR mutations while in September and October 2019, 6/18 (33%) cases of typhoid fever were caused by *S.* Typhi with QRDR mutations (Fisher’s exact test P= 4.679×10^-5^, Figure 2C). No other antibiotic had prescription trends that were similar to those of ciprofloxacin (Supplementary Figure 4).

Between September 2016 and August 2019 three different QRDR mutations were identified: GyrA S83F (n = 2), GyrA D87G (n = 1), and GyrB S464F (n = 1), while during September and October 2019, there were two different mutations identified - GyrB S464F (n = 5) and GyrA S83F (n = 1) (Figure 2D). Nine of the ten samples with QRDR mutations had a decreased ciprofloxacin susceptibility phenotype.

Regression modelling showed that each additional prescription of ciprofloxacin given to STRATAA and TyVAC participants was associated with a 4.2% increase in the relative risk of isolating *S.* Typhi with a QRDR mutation in the subsequent month (95% CI 1.8-7.0%, P = 0.0008, Table 1). The number of blood cultures taken was not significantly associated with the number of *S.* Typhi with QRDR mutations isolated in that month (OR = 1.00, 95% CI 1.00-1.00, P = 0.22).

**Table 1:** Results of epidemiological modelling to test the hypothesis that the proportion of *S.* Typhi isolated that had QRDR mutations were associated with the number of ciprofloxacin prescriptions in the preceding month. Model estimates have been exponentiated to yield odds ratios associated with lagged prescriptions.

| *Predictors* | *Odds Ratios* | *CI* | *p* |
| --- | --- | --- | --- |
| Total number of ciprofloxacin prescriptions in preceding month | 1.04 | 1.02 – 1.06 | 0.0008 |
| Total number of blood cultures | 1.00 | 1.00-1.00 | 0.224 |

### DCS S. Typhi isolated at Ndirande Health Centre in October-November 2019 formed two phylogenetically distinct groups and encoded two different QRDR mutations

We carried out a phylogenetic analysis of the 187 *S.* Typhi from different patients reported here, combined with 112 *S.* Typhi from Blantyre from previously published work (Feasey et al. 2015) (Figure 3). Five of the six *S.* Typhi isolates from Ndirande in Sept/Oct 2019 formed a subclade of near-identical isolates (median SNP pairwise distance = 0 SNPs) that shared the GyrB-S464F amino acid substitution. The sixth isolate was unrelated to the other 5 and encoded a GyrA-S83F amino acid substitution; its closest phylogenetic neighbour (4 SNPs) was from Ndirande Health Centre isolated in May 2019 that also encoded a GyrA S83F amino acid substitution.

**Figure 3:**
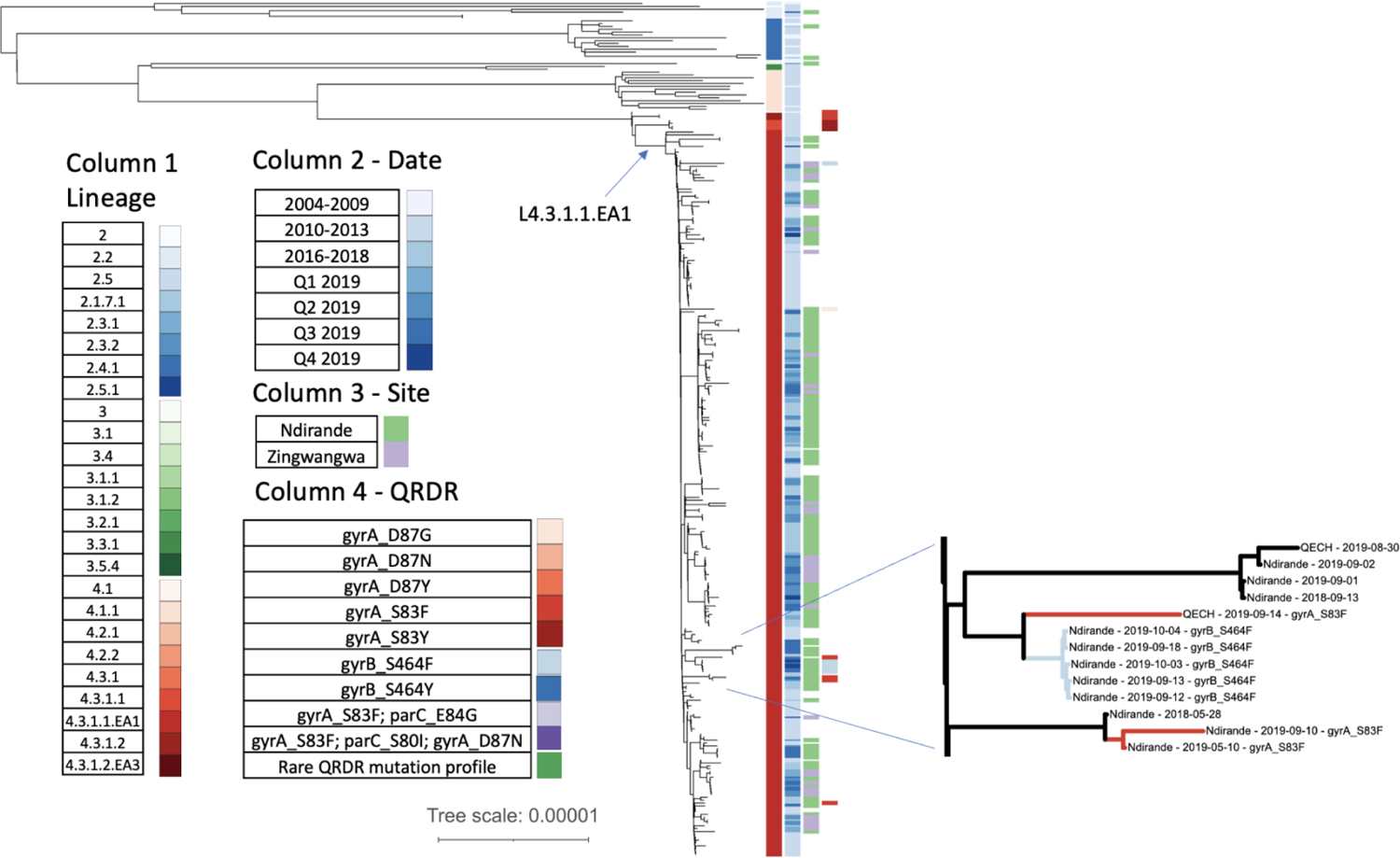
Phylogenetic tree of 299 S. Typhi from Blantyre (one genome per patient). The tree branches of the inset are coloured according to the QRDR mutation observed descending from that branch, the colour is the same as for column 4.

### DCS *S.* Typhi from Ndirande Health Centre was not the result of importation from outside Malawi

To investigate the hypothesis that the *S.* Typhi with QRDR mutations were imported from outside Blantyre, we derived a maximum likelihood phylogenetic tree of 299 Malawian genomes alongside an additional 1970 non-Malawian *S*. Typhi genomes from published studies. As the sub-clades of interest all belonged to H58, we present a sub-tree of 1168 H58 genomes (Figure 4) (Wong et al. 2015; Park et al. 2018; Mashe et al. 2021). Eighty-five per cent of Malawian genomes formed a paraphyletic clade belonging to L4.3.1.1.EA1 which also included 27 genomes from Zimbabwe, 4 genomes from South Africa, and 1 genome from an unspecified African country. The Zimbabwean genomes included isolates encoding both the QnrS1 plasmid-mediated quinolone resistance (PMQR) protein and QRDR mutations. An additional maximum likelihood phylogeny was constructed focussing on this sub-clade (Figure 5), which confirmed that the 27 Zimbabwean genomes formed a monophyletic clade and that no transmission of *S.* Typhi with QRDR mutations between Zimbabwe and Malawi was identified in this dataset. The most closely related genomes to the Blantyre *S.* Typhi with QRDR mutations were QRDR mutation negative *S.* Typhi from Blantyre.

**Figure 4:**
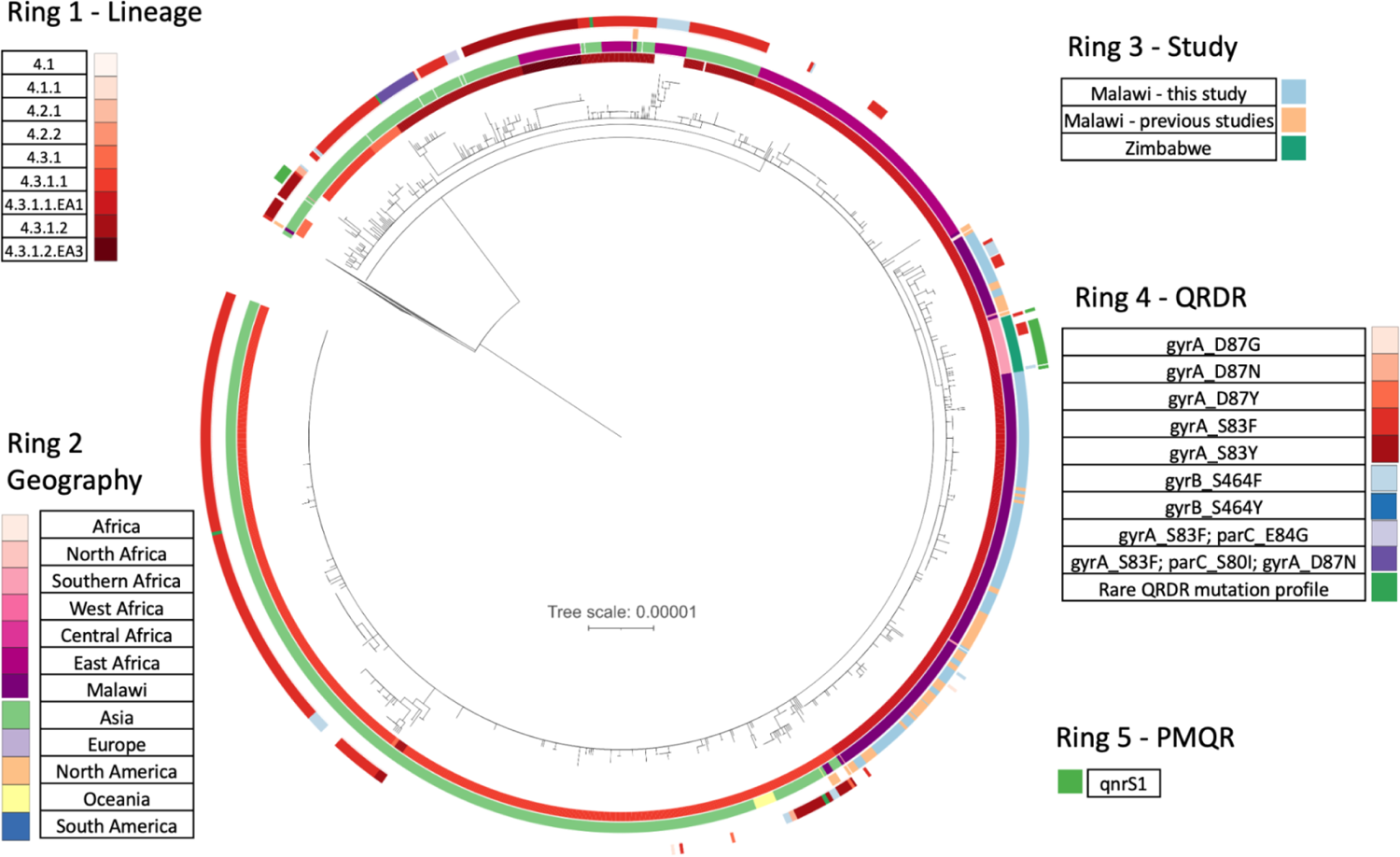
Phylogenetic tree of 1166 H58 S. Typhi genomes, placing those from Blantyre in the global context of other H58 samples

**Figure 5:**
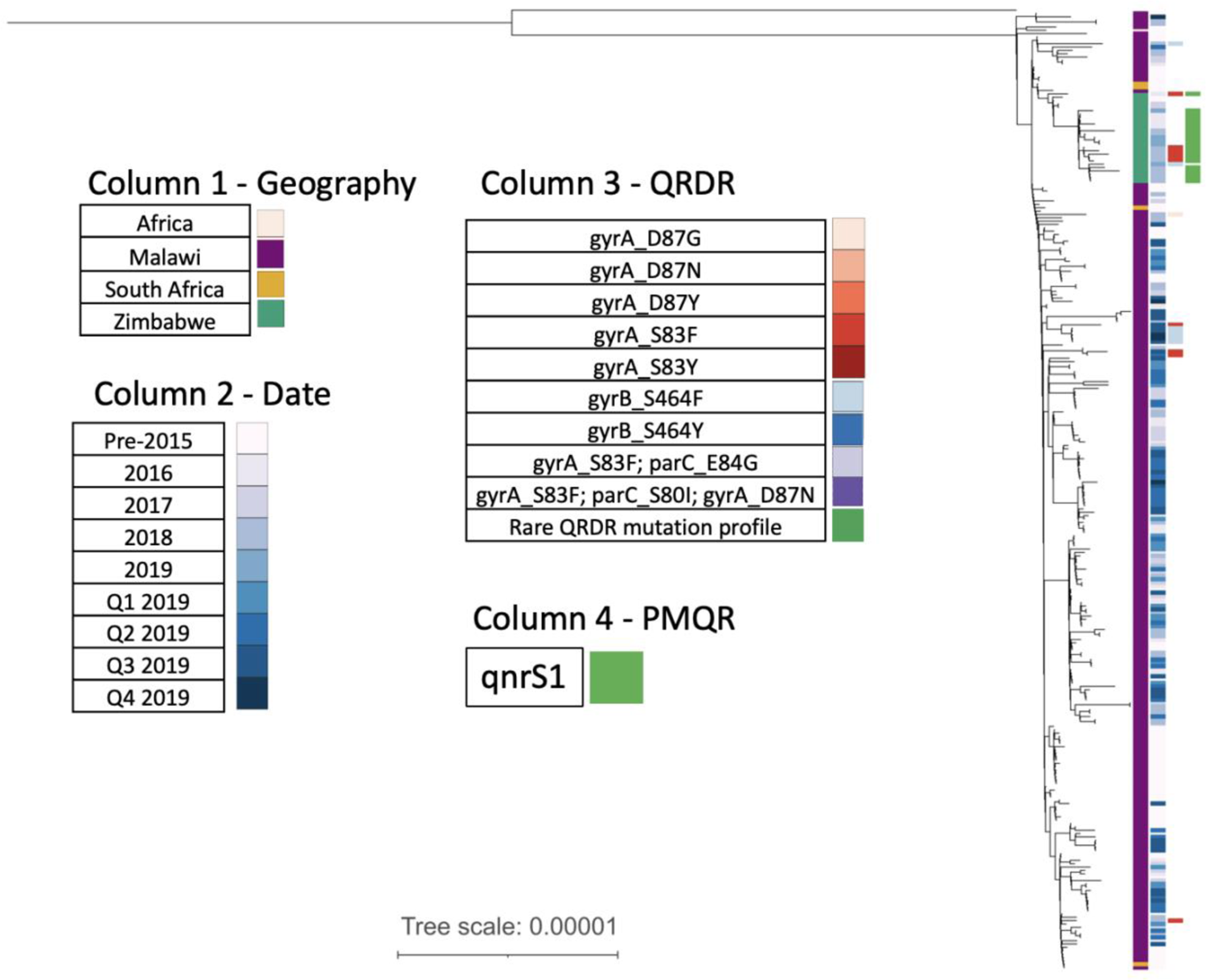
A phylogenetic tree of 286 genomes from the Malawian and Zimbabwean sub-clade. This clearly shows that the ciprofloxacin-resistant and DCS Typhi from Zimbabwe were genetically distinct from the Malawian DCS Typhi

## Discussion

### Statement of principal findings

We found a significant positive association between the proportion of *S.* Typhi encoding QRDR mutations and the number of courses of ciprofloxacin prescribed in the preceding month within a vaccine and epidemiological surveillance study in Blantyre, Malawi. Genomic evidence revealed that *S.* Typhi with QRDR mutations from Sept-Oct 2019 belonged to two different lineages, decreasing the probability of this association being driven by a stochastic event. Furthermore, placing our genomes into a broader genomic context revealed that the genomes most closely related to our DCS *S.* Typhi were ciprofloxacin-susceptible *S.* Typhi from Blantyre, and hence not the result of an introduction from another country.

### Strengths and weaknesses of the study

In our setting (district health centres in Blantyre, Malawi), clinical microbiology testing is not routinely available, and prescription information is not systematically captured. This is the case for most primary healthcare systems in countries in Sub-Saharan Africa, which is reflected in the paucity of primary literature on the association between antibiotic usage and antibiotic resistance in the region (Dromigny et al. 2005). Therefore, this study offered the valuable opportunity to identify and quantify factors associated with the occurrence of a WHO high-priority pathogen, fluoroquinolone-resistant *Salmonellae* in a setting with a high burden of typhoid fever (‘WHO Publishes List of Bacteria for Which New Antibiotics Are Urgently Needed’ 2017).

The use of genomics strengthened our analysis in multiple ways. Firstly, it enabled the highly reproducible identification of *S.* Typhi carrying QRDR mutations. Secondly, it enabled us to place our *S.* Typhi with QRDR mutations into the context of collections of *S.* Typhi genomes from Blantyre and internationally. This line of evidence revealed that the DCS *S.* Typhi emerged from Blantyre’s endemic *S.* Typhi population, rather than being imported from another country. Finally, it enabled us to identify that the *S.* Typhi with QRDR mutations identified in September and October 2019 belonged to two separate sub-clades with different QRDR mutations. While identifying two separate phylogenetic clades with QRDR mutations decreases the probability of the cluster resulting from a point-source outbreak, 5 of the 6 *S.* Typhi with QRDR mutations were monophyletic and very closely related (median pairwise distance was 0 SNPs). However, this is difficult to interpret for *S.* Typhi, where isolates obtained years apart and across wide geographic spans can be 0 SNPs distant (Dyson et al. 2023).

With our ecological study design (i.e. we looked at population-level emergence of resistance associated with AMU, not individual-level emergence), we cannot definitively prove causality in the relationship between ciprofloxacin use and QRDR emergence. However, it is worthwhile to consider the plausibility of forward causality (i.e. an increase in ciprofloxacin prescriptions caused more *S.* Typhi with QRDR mutations) and reverse causality (i.e. an increase in *S.* Typhi with QRDR mutations caused more ciprofloxacin prescriptions). Forward causality is biologically plausible and the ciprofloxacin prescriptions given in the prior month were significantly associated with the proportion of *S.* Typhi with QRDR mutations, a temporal relationship which supports the forward causal relationship. Reverse causality could plausibly occur when prescriptions are made with reference to local AMR trends, but it is not plausible that the response to an increase in DCS *S.* Typhi would be to increase ciprofloxacin prescriptions. Therefore, the forward causal relationship seems the most likely explanation, although there may be sources of bias that have not been accounted for (e.g. ciprofloxacin usage outside the study). Another disadvantage of the ecological study design is that we did not observe the emergence of resistance in patients treated with ciprofloxacin because samples were taken for blood culture before the prescribed antibiotics were taken, although it is unclear whether we would have observed that as drug resistance in *S.* Typhi may be due to the bystander effect (Morley, Woods, and Read 2019).

The majority of human antibiotic use in Africa is not by prescription from a health worker (Morgan et al. 2011; Auta et al. 2019). Although reports from Malawi suggest that a higher than average 64% of antibiotics are prescribed by health facilities (Dixon et al. 2021), this still leaves a substantial fraction of people who obtain antibiotics informally. As we only captured ciprofloxacin prescriptions given as part of the STRATAA and TyVAC studies, there is the possibility of bias due to the usage of ciprofloxacin outside of our study. However, in terms of informal use, ciprofloxacin is not commonly used as a non-prescription antibiotic in Malawi, with only 2-3.2% of people reporting using oral ciprofloxacin “frequently”, compared with 68% and 36% of people reporting frequent use of cotrimoxazole or amoxicillin respectively (Dixon et al. 2021; MacPherson et al. 2022). In terms of prescribed ciprofloxacin that was not given as part of our study, we have no way of reliably quantifying this either within the areas we were recruiting in, or in Blantyre more broadly.

One major weakness of our study is that we only observed a relatively small number of *S.* Typhi with QRDR mutations, which limits the strength of our findings. There is also no evidence from the hospital-based surveillance (Figure 1) of a broader increase in DCS *S.* Typhi, although interpretation of this is difficult, as the failure of the DCS *S.* Typhi to fix in the population could also be linked to the low level of ciprofloxacin usage in Malawi. Furthermore, there was a decrease in the number of *S.* Typhi-positive blood cultures detected by hospital based surveillance following the emergence of SARS-CoV-2, probably driven by a reduction in hospital attendance because of fear of the virus.

Models were not adjusted for any co-variables because our outcome measure was the proportion of *S.* Typhi that encoded QRDR mutations, and our methodology for identifying these was consistent across the study period. If we had used the number of *S.* Typhi with QRDR mutations then we would also need to take into account the number of blood cultures carried out and the subtly different sampling frames of the two studies.

### Our findings in relation to other studies

Our modelling revealed a significant positive association between ciprofloxacin prescriptions and the proportion of *S.* Typhi with QRDR mutations in the following month. This short lag time is in line with what other studies have found for the relationship between ciprofloxacin prescriptions and fluoroquinolone resistance (Vernaz et al. 2011; Sun et al. 2022; Gottesman et al. 2009). However, much of the work in the literature other work has been done for *E. coli* rather than *Salmonella* infections, so we cannot be certain of its relevance. Uncontrolled usage of antibiotics has long been blamed for the emergence of resistance to fluoroquinolones and other antibiotics in *S.* Typhi in high infectious disease burden settings (Britto et al. 2018; Wain et al. 1997), but empirical demonstrations of this relationship have been lacking. However, there have been many demonstrations of the association between ciprofloxacin usage and resistance in Enterobacteriaceae in high-income countries - in hospitals (I. Willemsen et al. 2009; Cusini et al. 2018; Batard et al. 2013; Ina Willemsen et al. 2010), communities (Vernaz et al. 2011; Bryce, Costelloe, et al. 2016; MacDougall et al. 2005; Bryce, Hay, et al. 2016; Olesen et al. 2018), and whole countries (van de Sande-Bruinsma et al. 2008). These studies show similar effect sizes to our analysis (e.g. 1% increase in ciprofloxacin prescriptions led to a 0.97% increase in ciprofloxacin-resistant isolates 1 month later (Vernaz et al. 2011)). Studies that showed that fluoroquinolone stewardship interventions reduced either the incidence or the rate of increase of fluoroquinolone resistance demonstrate that this is likely a causal association in some circumstances (Gottesman et al. 2009; Ina Willemsen et al. 2010). Our study contributes to this evidence base by demonstrating the association between prescribed antibiotics and *S.* Typhi with QRDR mutations in a community setting in sub-Saharan Africa, where there are only a limited number of existing studies (Dromigny et al. 2005). However, it is weaker than other studies on this topic because we took an ecological approach, as opposed to demonstrating an individual-level effect (Bryce, Hay, et al. 2016; Bryce, Costelloe, et al. 2016).

### Implications for clinicians and policymakers

We have identified an association between ciprofloxacin usage and the proportion of *S.* Typhi with QRDR mutations. If this relationship is causal, then there are important policy implications.

Only around 1% of febrile patients in Blantyre have typhoid fever (Patel et al. 2021; Meiring et al. 2021). Therefore, very few of the courses of ciprofloxacin prescribed as part of our study were given to people for whom *S.* Typhi was the cause of their fever. There are two niches in which there is likely more *S.* Typhi exposure to ciprofloxacin than in typhoid fever patients. The first niche is in chronic carriers or asymptomatically infected people who were treated with ciprofloxacin because of a fever with a non-typhoidal cause. The sero-incidence of *S.* Typhi exposures/infections in Blantyre has been estimated as 43 times higher than the observed incidence (Meiring et al. 2021). We speculate that there are likely to be more people presenting with fever who have *S.* Typhi as an asymptomatic or sub-clinical infection than those for whom *S.* Typhi is the cause of their fever. Furthermore, it is estimated that around 2-5% of the population in endemic regions are chronic carriers of *S.* Typhi (Gunn et al. 2014). This represents a high burden of asymptomatic, sub-clinical, or chronic infections which could be evolving in response to ciprofloxacin given because of fever induced by a different cause. The second niche is in the environment e.g. water sources (Karkey et al. 2016). Between 60-87% of ciprofloxacin administered to people is excreted unmetabolised via the urine or faeces (Girardi et al. 2011). Levels of ciprofloxacin observed in the environment have been shown to inhibit bacterial growth (Girardi et al. 2011). We believe it is possible that the increased number of ciprofloxacin prescriptions would lead to more contamination of the environment with ciprofloxacin, which could select for QRDR mutations. However, we expect that this indirect route of ciprofloxacin exposure would have a longer lag between ciprofloxacin prescriptions and the emergence of non-susceptibility in the human population than we observed in this study. Regardless of the niche in which the exposure occurred, an accurate point-of-care diagnostic for typhoid fever would help to reduce overall ciprofloxacin usage in Malawi by targeting treatment to people whose fever is caused by *S.* Typhi. We hypothesise that this reduced overall usage would reduce the chance of non-susceptibility emerging.

More direct modification of AMU policies, rather than just better diagnosis leading to fewer prescriptions, is another option that could potentially reduce the emergence of resistance. One AMU policy that could be modified is to increase the dose of ciprofloxacin given, to achieve a “mutant prevention concentration”, although this has not shown promise in the limited empirical studies carried out thus far (Fantin et al. 2009). Another antimicrobial usage policy that could be considered is a switch to combination therapy for suspected typhoid fever (Zmora et al. 2018; Giri et al. 2021), although without improved diagnostics this might just serve to increase the overall antibiotic pressure.

It is notable that in both Zimbabwe and Malawi in the late 2010s, there have been outbreaks of ciprofloxacin non-susceptible *S.* Typhi that emerged from the local population of *S.* Typhi, rather than being imported from another setting (Mashe et al. 2021; Thilliez et al. 2022). The emergence of non-susceptibility due to local AMU practices empowers local stakeholders to advocate for the tools to enable better practices. If the DCS *S*. Typhi were due to importations, it is very hard to prevent these kinds of events, but non-susceptibility arising in the endemic bacterial population due to local practices can potentially be modified by changes in local practice.

Ultimately, reducing the burden of disease through systemic measures such as the introduction of efficacious vaccines into high-burden settings and WASH interventions are likely to be the most comprehensive solutions to preventing the emergence of DCS *S.* Typhi (Patel et al. 2021; Qadri et al. 2021; Shakya et al. 2021). Indeed, typhoid conjugate vaccines were introduced into Zimbabwe in response to an outbreak of typhoid there in 2018 that included a large proportion of ciprofloxacin non-susceptible isolates (‘Stories from the Field: How Vaccines Can Help to Prevent Antibiotic Resistance - Zimbabwe’s Response to Drug-Resistant Outbreaks of Typhoid and Cholera’ n.d.; Mashe et al. 2021; N’cho et al. 2019).

## Data Availability

Phylogenetic trees and annotation files are available from FigShare at the following link https://figshare.com/projects/Fluoroquinolone_resistance_Salmonella_Typhi_in_Blantyre_Malawi/157785. Code to reproduce the statistical analysis and generate the plots can be accessed from https://github.com/flashton2003/blantyre_qrdr_typhi. Genome accessions are available in Supplementary Table 1.

## Acknowledgements

We would like to acknowledge all the participants in the STRATAA and TyVAC studies for their willingness to participate in this research, and also all the team and consortium members for their contributions. We would like to acknowledge the contribution of Sithembile Bilima and the other members of the MLW Core laboratories staff for their valuable contributions to this work. We thank the Wellcome Sanger Institute core laboraroty and pathogen informatics teams for their assistance.

## Consortium members

STRATAA consortium members - Happy Chimphako Banda^1^, Prasanta Kumar Biswas^2^, Md. Amiruli Islam Bhuiyan^2^, Christoph Blohmke^3^, Thomas C. Darton^3^, Christiane Dolecek^4^, Sabina Dongol^5^, Yama Farooq^3^, Jennifer Hill^3^, Nhu Tran Hoang6, Tikhala Makhaza Jere^1^, Maurice Mbewe^1^, Harrison Msuku^1^, Tran Vu Thieu Nga^6^, Rose Nkhata^1^, Sadia Isfat Ara Rahman^2^, Nazia Rahman^2^, Neil J. Saad^7^, Trinh Van Tan^6^, Deus Thindwa^1^, Merryn Voysey^3^, Richard Wachepa^1^

## Affiliations

1. Malawi-Liverpool Wellcome Programme, Blantyre, Malawi
2. International Centre for Diarrhoeal Disease Research, Dhaka, Bangladesh
3. Oxford Vaccine Group, Department of Paediatrics, University of Oxford, and the NIHR Oxford Biomedical Research Centre, Oxford, United Kingdom
4. Nuffield Department of Medicine, Centre for Tropical Medicine and Global Health, University of Oxford, Oxford, UK
5. Oxford University Clinical Research Unit, Patan Academy of Health Sciences, Kathmandu, Nepal
6. The Hospital for Tropical Diseases, Wellcome Trust Major Overseas Programme, Oxford University Clinical Research Unit, Ho Chi Minh City, Vietnam
7. Department of Epidemiology of Microbial Diseases and the Public Health Modeling Unit, Yale School of Public Health, Yale University, New Haven, CT, USA

