## Supplementary Figures for "The rapid emergence of *Salmonella* Typhi with decreased ciprofloxacin susceptibility following an increase in ciprofloxacin prescriptions in Blantyre, Malawi"

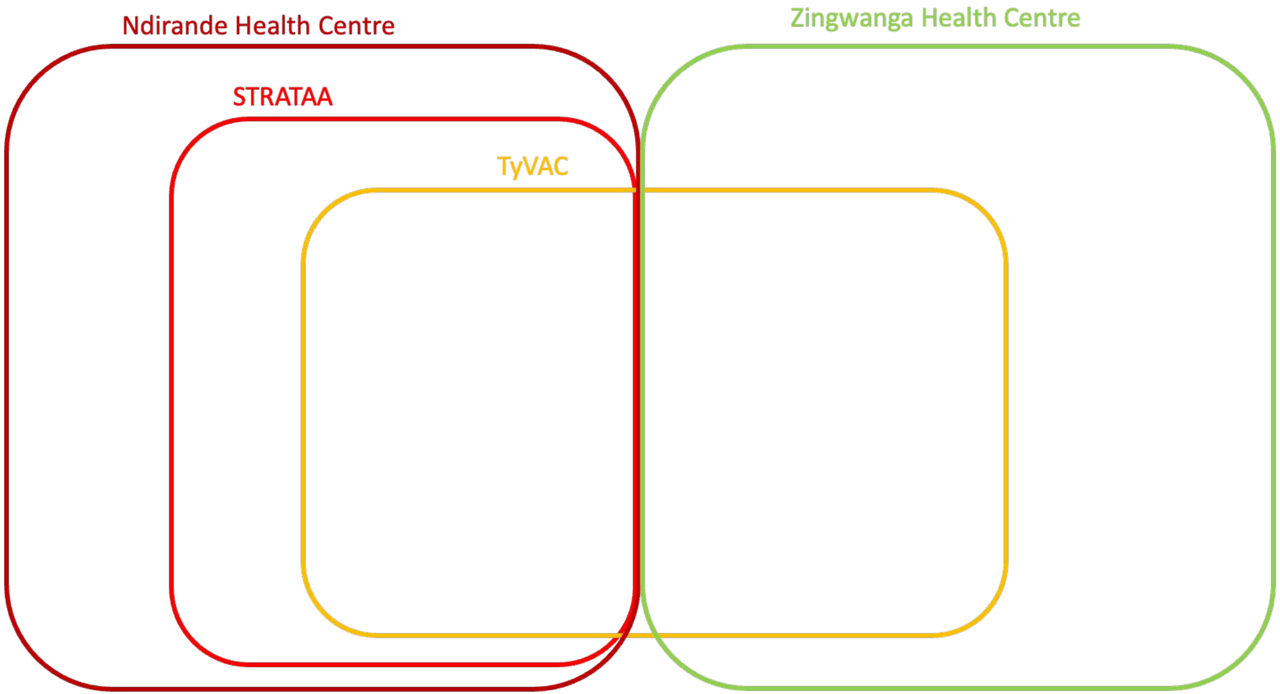


Supplementary Figure 1: A schematic diagram demonstrating the relationship between the STRATAA and TyVAC cohorts, and their recruitment from Ndirande and Zingwangwa. TyVAC recruited from both Ndirande and Zingwanga, while STRATAA only recruited from Ndirande. All TyVAC participants from Ndirande were also enrolled in STRATAA. TyVAC participants from Ndirande were only those enrolled in the typhoid conjugate vaccine study, while all residents were eligible for enrollment in STRATAA (assuming they met the fever criteria etc).

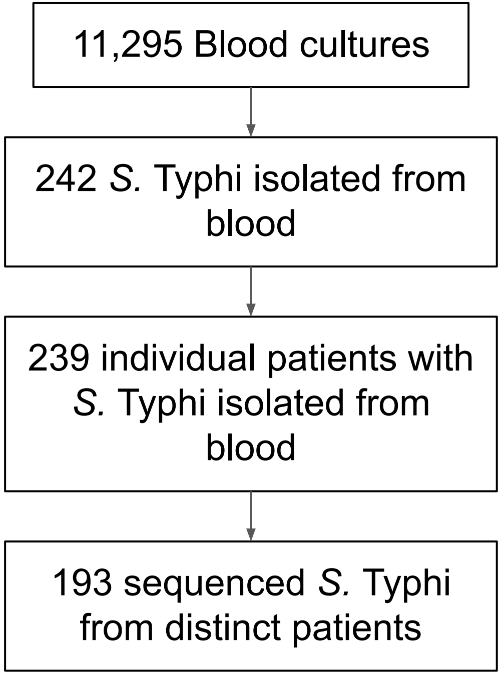


Supplementary Figure 2: STROBE diagram of blood cultures done, *S.* Typhi isolated and *S.* Typhi sequenced as part of the STRATAA and TyVAC studies

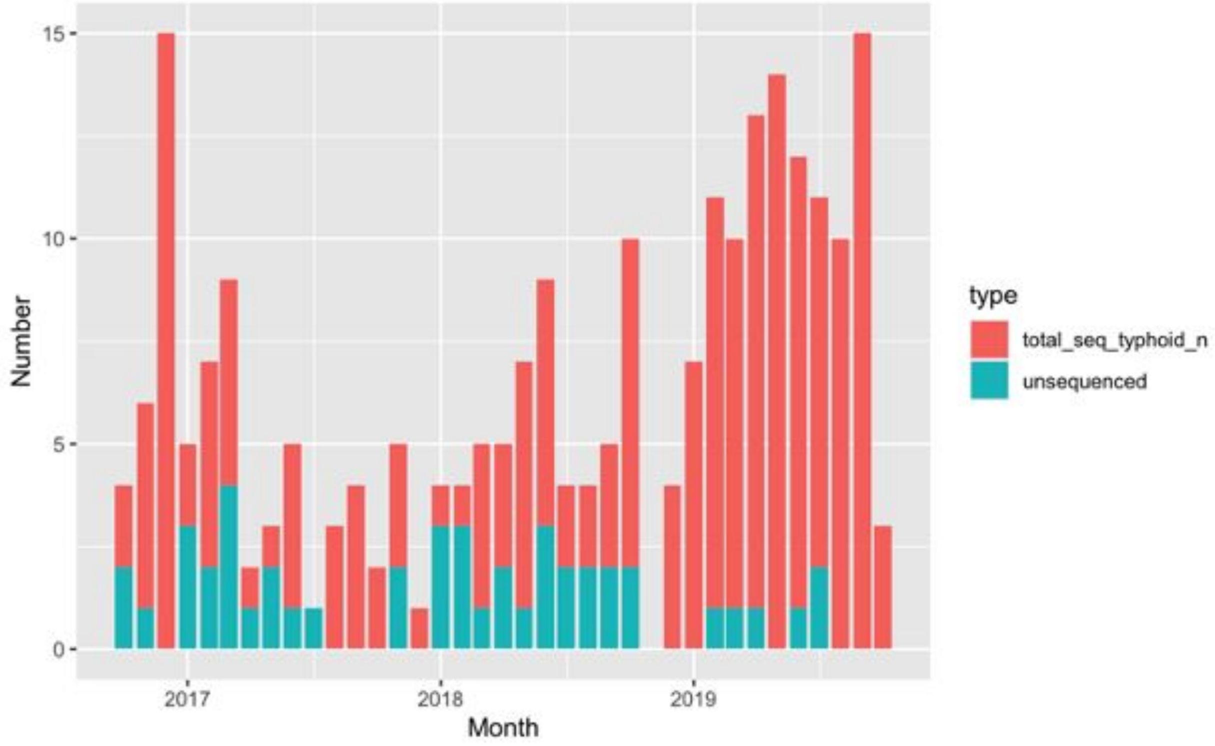


Supplementary Figure 3: The number of patients with sequenced and unsequenced *S.* Typhi isolates across the recruitment period

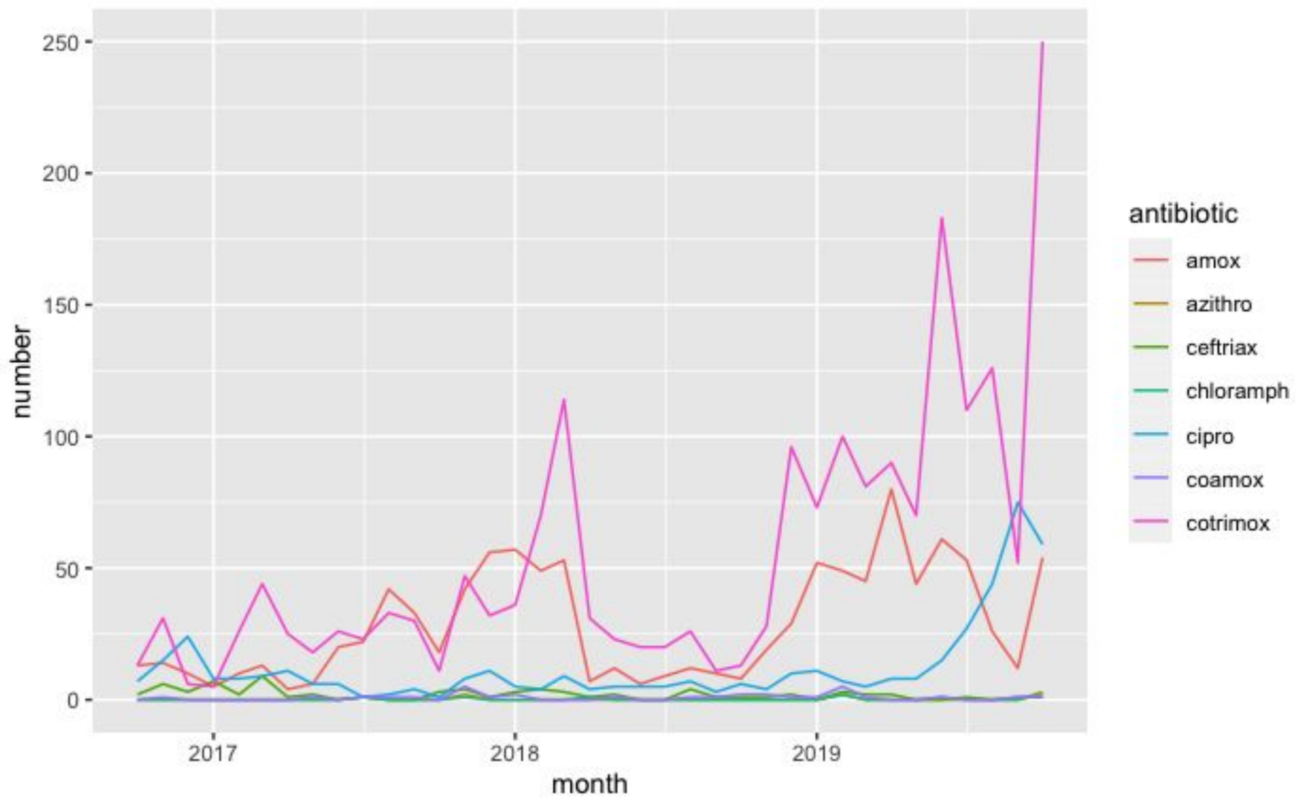


Supplementary Figure 4: Monthly prescriptions of 7 antibiotics to STRATAA participants
